# The Inflation Reduction Act’s Impact upon Early-stage Venture Capital Investments

**DOI:** 10.1101/2025.01.07.25320113

**Authors:** Duane G. Schulthess, Gwen O’Loughlin, Madeline Askeland, Daniel Gassull, Harry P. Bowen

## Abstract

**Background:** The Congressional Budget Office has stated there is no evidence of a systematic decrease in the percentage of venture capital flowing to pharmaceutical companies since IRA’s passage. This was echoed in Prof. Rita Conti’s September 17, 2024, Senate Finance Committee testimony.

**Methods:** To test the IRA’s impacts on early-stage investments targeting therapeutics for the Medicare-aged population, a longitudinal dataset of commercially sponsored clinical trials by companies with a market valuation < $2 billion was obtained from the BioMedTracker database from January 1, 2018, to May 6, 2024. These trials were filtered to match early-stage investments to lead assets undergoing clinical development.

**Results:** From 161 lead assets with 897 investments, we find the aggregated total into large molecules in 2024 was 10 times larger than that for small molecules, which underwent a 70% decline after passage of the IRA. Individual investments made into small molecules decline by minus one-half as exposure to the Medicare-aged population increases after the passage of the IRA (p < 0.0018). Testing large molecule investments by their exposure to Medicare post IRA’s passage is statistically inconclusive.

**Research Conclusions:** This study presents evidence of a decline in the development of new therapies targeting the Medicare-aged population since the passage of the IRA. If these impacts were due to the economic downturn post-pandemic, we would observe statistically similar results in both large and small molecules. However, the results by molecule type diverge. Investors perceive large molecules to be of a lower investment risk relative to small molecules after IRA’s passage.

## Introduction

Introduced as The Build Back Better Act on September 27, 2021, the Inflation Reduction Act (IRA) was signed into law by the Biden Administration on August 16, 2022, for the first time, “allowing Medicare to negotiate prescription drug costs.” (1).

The Congressional Budget Office (CBO), in their scoring of the IRA, reported that it would reduce direct spending on drugs by $249 billion. CBO further stated that the impact would be limited as “the number of drugs that would be introduced to the U.S. market would be reduced by about two over the 2023-2032 period, about five over the subsequent decade, and about eight over the decade after that.” (2).

Many leading academics have echoed this belief that the IRA is relatively benign for the U.S. biopharmaceutical innovation ecosystem. In a September 17, 2024, Senate Finance Committee hearing, (3) Prof. Rena M. Conti presented research stating they “found little evidence the IRA has resulted in a meaningful decrease in the level of venture capital (VC) investment in new drug development.” (4). Prof. Conti also said that “Late-stage private company and public equity valuations, IPOs, follow-on offerings, and mergers and acquisitions in biopharma largely held steady in the 18 months after passage of the IRA and [that such investments] have had a positive start in 2024.” (5).

The IRA allows the Center for Medicare and Medicaid Services (CMS) to set prices for the top 20 drugs by spending beginning year 9 for small molecules and year 13 for large molecules, creating a two-tiered reimbursement system. The challenge for investors then, particularly those operating at the early stages of drug development, is that the IRA’s provisions create specifically divergent financial disincentives for large and small molecule therapies, including market risks as measured by a disease’s prevalence in the Medicare-aged population.

The research cited above suggests the IRA poses little risk to biopharmaceutical development. However, the methodological approach taken in these studies only examines the IRA’s impacts in the aggregate, i.e., only the average impacts. They do not segment by large or small molecules, measure a specific indication’s exposure to the Medicare-aged population, or differentiate late-stage phase III research from earlier stages in smaller, highly innovative firms where the IRA’s impacts are more likely to first appear due to shifts in VC or angel investing behavior.

There is evidence that VCs and developers are responding to the IRA’s disincentives. For example, on February 9, 2024, Suneet Varma, commercial president of Pfizer Oncology, stated, “Biologics [represent] a more durable revenue potential based on several factors, including differentiated access and affordability to the patient, IRA considerations and patent expiration timeline.” (6) Similarly, venture capitalist Peter Kolchinsky stated, “We’ve told our companies… stay away from any disease of aging where you’re going to be heavily dependent on Medicare.” (7).

In addition to potentially missing these changes in investor behavior, the aggregated approach of prior research is also likely to underestimate the IRA’s microeconomic impacts by not capturing the relationship between Biopharmaceutical profits and research & development (R&D). Specifically, as recently highlighted by Chandra et al., “The top 20 companies by revenue (accounting for 71% of total revenue) contribute 50% of R&D investment.” (8). This implies that the Biopharmaceutical sector’s profits and R&D spending follow the Pareto principle, whereby 20% of successful companies fund an outsize portion of the entire R&D ecosystem (9). If true, we would expect the first responses to the law’s disincentives to be observed among early-stage investors and drug developers who would avoid future exposure to the IRA’s provisions by altering their exposure to the Medicare-aged population, particularly in small molecules.

Vogel et al. said, “Capital already committed to biopharmaceutical VC funds will be deployed even if return expectations have shifted. This is not necessarily true for VC funds seeking new capital.” (10). Consistent with this remark, this paper uses both descriptive statistics and formal statistical tests to detect statistically significant changes in investing behavior concerning early-stage funding of large and small molecules and in developing assets with high exposure to the Medicare-aged population and therefore IRA price setting. The analysis is conducted using a purposely constructed dataset of recent investments made by early-stage VC and angel investors, segmented at the indication and molecule level.

## Materials and Methods

Our dataset was built by extracting data from BioMedTracker (11) and ClinicalTrials.gov for the period January 1, 2018 to May 6, 2024 on U.S. companies with a market capitalization/valuation of less than our equal to $2 billion, which is the standard forward-looking risk-weighted net present value of a successful firm at the time of a therapeutic product’s FDA submission for marketing authorization (12). The $2 billion cutoff was chosen to ensure our cohort would focus on small to mid-sized firms and include drugs under clinical development requiring early-stage funding for therapies that would potentially be subjected to IRA negotiations if eventually commercialized. This cohort comprised 1,137 clinical trials.

As many molecules undergoing clinical trials will be developed for multiple indications simultaneously, and drugs often change names or are redeveloped under altered formulations, each of the 1,137 individual clinical trials was researched on a company-by-company level to determine which of these programs was functioning as a given company’s “lead” asset. To identify a company’s lead asset, all clinical trials for which the FDA disclosed the company as the lead sponsor were searched to determine which asset was at the highest phase of development. These registered trials were then cross-referenced with any forward-looking statements filed with Securities and Exchange Commission’s (SEC) audited Federal filings, publicly circulated press releases or statements made to the media. This process yielded 228 lead assets, with 897 individual investments. These lead assets were then segmented by indication and clinical trial phase to focus on only those lead assets in either phase I or II clinical trials, yielding 161 early-stage lead assets.

In several cases, our lead asset selection process found that a molecule was being developed for either an accelerated approval pathway, an orphan pathway, or both simultaneously and with varying indications. In these cases, we choose the indication with the largest potential market size as the company’s leading asset.

There were also several cases of an investigational therapy that had undergone multiple failed phase II clinical trials and had then been registered in a clinical trial addressing the COVID-19 pandemic. We regarded these trials as merely opportunistic and opted to use the earlier, non-COVID-19 indication as the lead asset, except in those rare cases where investors had shown significant financial interest in the COVID-19 trial where it was legitimately a company’s lead asset.

Once we identified lead assets by company, we executed a forensic audit of all identifiable investment activity for each lead asset between January 1, 2018, and August 16, 2024, using the combined resources of Pitchbook, BioMedTracker, SEC filings, press releases, and published annual corporate reports. Our investment criteria focused on capital raised from VC, angel, equity, partnering, initial public offerings (IPO), and licensing activity. Debt assumed by the developing company is excluded. Where a clear audit trail of investment or ownership was not possible, those companies and developments were excluded from our analysis.

Finally, our analysis focuses on Type-1 novel FDA-approved therapies. Follow-on indications or post-approved combination therapies are excluded. The investment data are measured in July 2024 constant dollars.

## Results

### Drug Development Impacts

Figure 1 shows the evolution in clinical trial launches by our cohort of companies between 2018 and 2023, which reveals a 35% decline in clinical trial starts after the passage of the IRA in September 2021. A regression test for the difference in the mean value of monthly clinical trials begins in the years before and after the passage of the IRA indicates a statistically significant difference (p ≤ 0.0179), thus affirming a statistically significant reduction in the post-IRA median number of trial launches, which is also validated by recently published research by the National Pharmaceutical Council (13).

**Figure 1.**
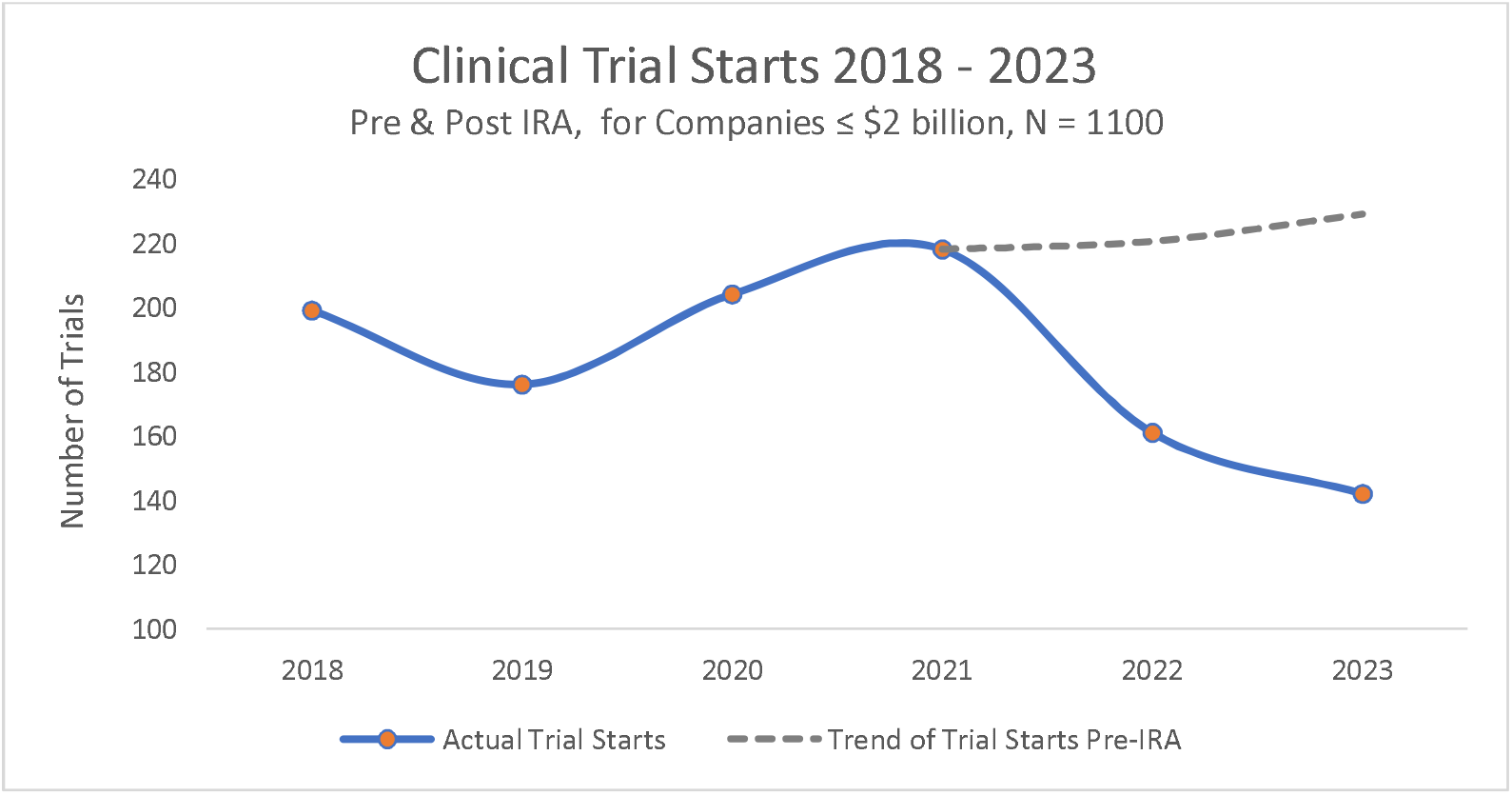
Number of clinical trials launched between 2018 and 2023 shows a statistically significant (p ≤ 0.0179) decline of 35% after introduction of the IRA on September 27^th^, 2021. Data cohort is 1,130 phase I and II clinical trials extracted from BioMedTracker for companies ≤ $2 billion valuation.

### Impacts on Investor Behavior – Size of Investments

Figure 2 shows the total annual investments into our cohort of 161 lead assets from 2018 to 2024 segmented by large and small molecules. As evident in Figure 2, aggregate investments in large molecule assets underwent a significant decline starting in 2021, coinciding with the passage of the IRA, but thereafter rose substantially, with aggregate large molecule investments being 10 times larger than those in small molecules by 2024.

**Figure 2.**
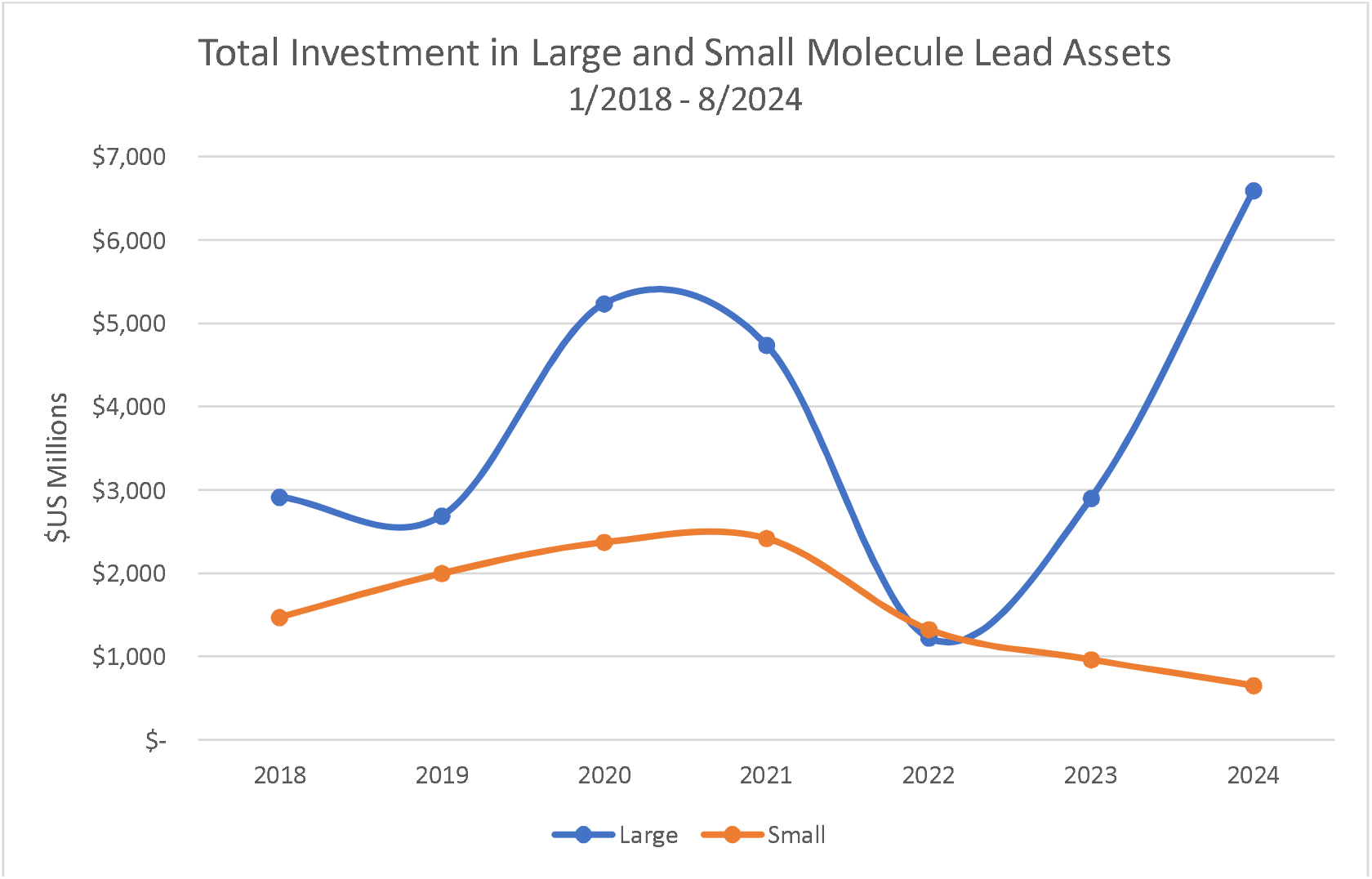
Graph showing large shift in early-stage (phase I and II) investments into small (MNE) to large (Biologics) molecule lead assets. Data for U.S. companies with a market value under $2 billion, values in 2024 constant dollars, 1/1/2018 – 8/16/2024.

Conversely, Figure 2 shows that aggregate investments in small molecules underwent a more than 70% decline after passage of the IRA, which axiomatically demonstrates a marked decrease in lead asset investments into small molecules relative to those into large molecules.

Figure 3 provides further evidence of the decline in small molecule investments following introduction of the IRA by showing the distribution of the size of investments into small molecules for the periods before and after passage of the IRA. Based on these data, the Kruskal–Wallis test indicates a significant difference (p < 0.0099) in the median size of small molecule investments pre- and post-IRA, affirming the significant post-IRA decline in small molecule investments observed in Figure 2.

**Figure 3.**
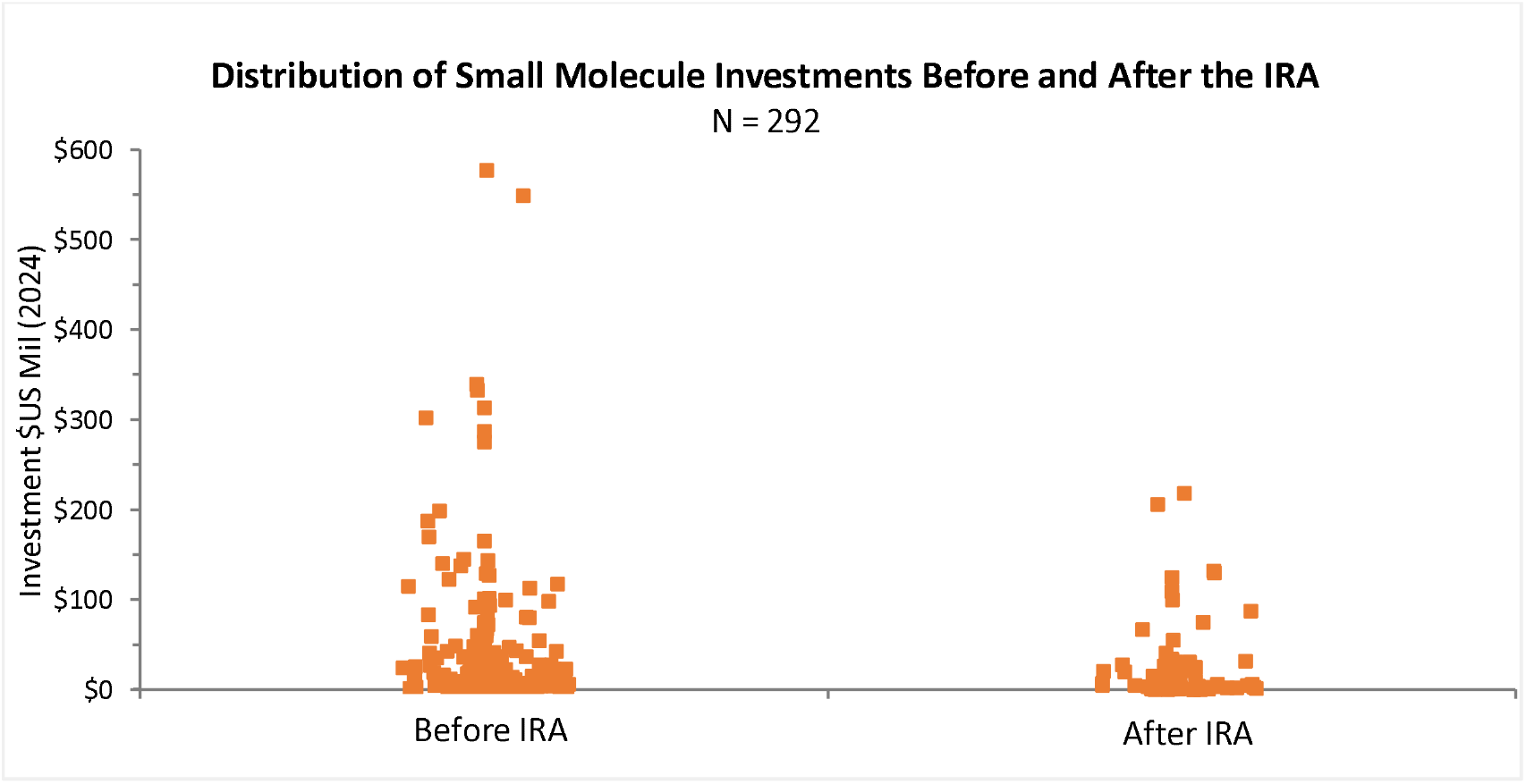
Dot plots show distributions, before and after IRA passage, of the size of early stage (phase I and II) investments into small molecule lead assets. Data are for 1/1/2018 – 8/16/2024 on U.S. companies with a market value ≤ $2 billion, investment values in 2024 constant dollars.

For large molecules, the Kruskal–Wallis test indicates a statistically significant decline (p < 0.023) in the median size of large molecule investments before and after the IRA (Figure 4). The previously observed increase in the total funding of large molecules in the aggregate (Figure 2) was due to three large post-IRA investments, each over $1 billion. Of note is these three post-IRA investments were for indications with an average exposure to the Medicare-aged population of 43%, a value in the lowest quartile of our measure of an indication’s exposure to the Medicare-aged population and hence also with the lowest exposure to the IRA’s provisions (14).

**Figure 4.**
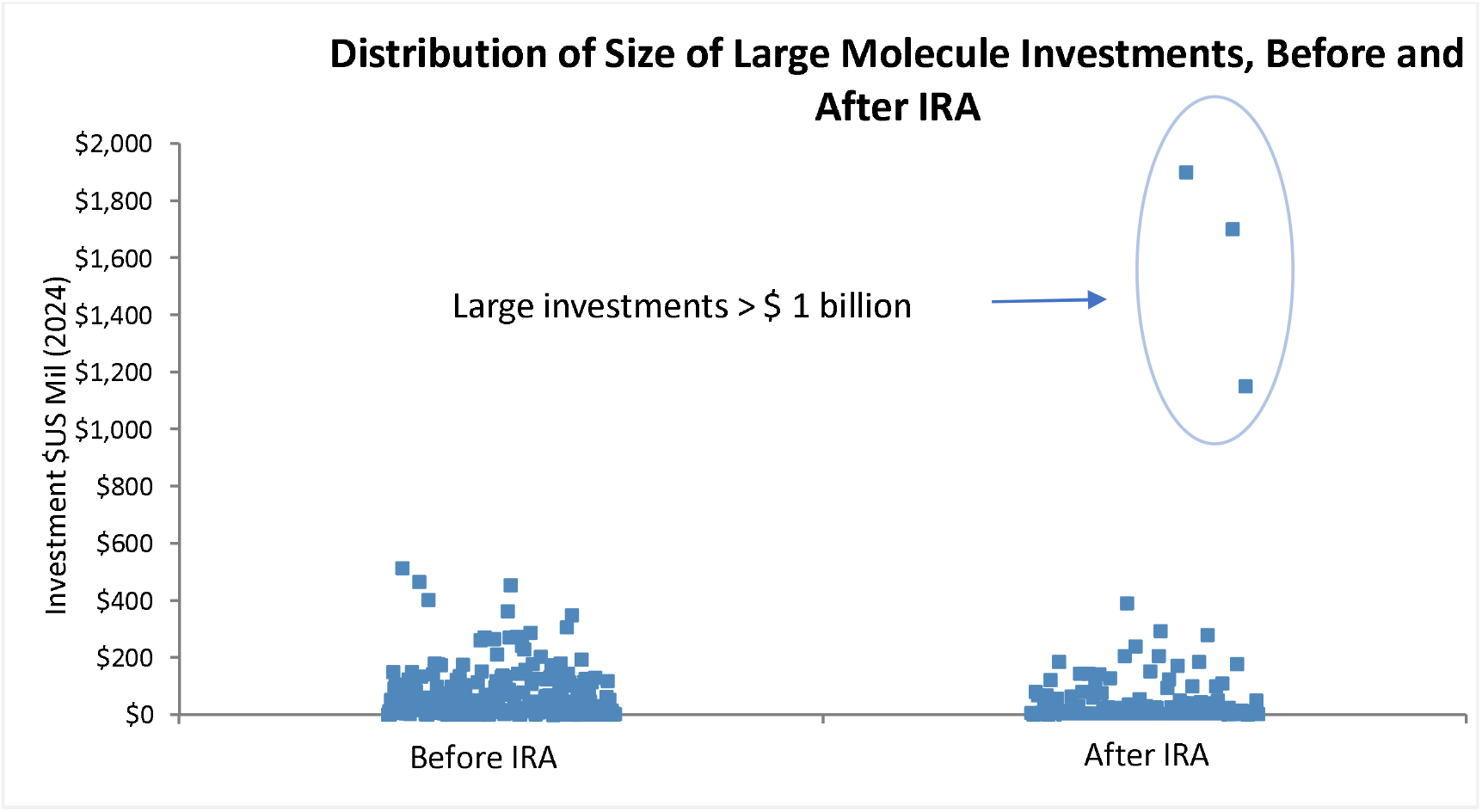
Dot plots show distributions before and after passage of the IRA of the size of early stage (phase I and II) investments into 327 large molecule lead assets. Data are for 1/1/2018 – 8/16/2024 on U.S. companies with a market value under $2 billion, investment values in 2024 constant dollars. Investment outliers are circled.

### Impacts on Investor Behavior – Exposure to the Medicare-aged Population

To further assess the evolution of early-stage investment behavior regarding lead assets with higher or lower exposure to the Medicare-aged population, our 161 lead assets were filtered to focus only on those indications whose measured exposure to the Medicare-aged population exceeded its median value (59%) in our cohort. Within this sample, the median size of the 242 investments into these 161 lead assets shows a statistically significant (p ≤ 0.0057) decline of 67% from its pre-IRA value (Table 1).

**Table 1.** Indications in our cohort with a high exposure to Medicare reimbursement for U.S. companies with less than $2 billion market value. Investments measured in 2024 constant dollars are for phase I and II from 1/1/2018 – 8/16/2024. Using the Kruskal–Wallis test, the observed difference in the median investment size before and after the IRA is statistically significant (p ≤ 0. 0057).

| Indication | Number of Investments | Prevalence in Medicare-Aged Population | Investments (\$ bil.) | |
| --- | --- | --- | --- | --- |
|  |  |  | Before IRA | After IRA |
| Non-Small Cell Lung Cancer (NSCLC) | 36 | 72% | 2,262 | 823 |
| Alzheimer's Disease (AD) | 30 | 95% | 189 | 97 |
| Multiple Myeloma (MM) | 26 | 65% | 2,250 | 529 |
| Acute Myelogenous Leukemia (AML) | 25 | 61% | 1,036 | 535 |
| Pancreatic Cancer | 13 | 69% | 270 | 299 |
| Prostate Cancer | 11 | 63% | 1,721 | 43 |
| Head and Neck Cancer | 11 | 70% | 167 | 14 |
| Gastric Cancer | 10 | 60% | 369 | 40 |
| Wet AMD | 8 | 90% | 200 | 626 |
| Non-Tuberculous Mycobacteria (NTM) | 8 | 60% | 178 | 85 |
| Acute Renal Failure (ARF) | 7 | 67% | 176 | 0 |
| Cartilage and Joint Repair | 7 | 75% | 63 | 0 |
| Idiopathic Pulmonary Fibrosis (IPF) | 6 | 90% | 110 | 37 |
| Rheumatoid Arthritis (RA) | 6 | 61% | 44 | 0 |
| Amyotrophic Lateral Sclerosis (ALS) | 6 | 64% | 68 | 204 |
| Acute Decompensated Heart Failure (Acute HFrEF) | 5 | 80% | 175 | 16 |
| Dementia | 4 | 95% | 716 | 0 |
| Renal Cell Cancer (RCC) | 3 | 71% | 83 | 0 |
| Stroke Prevention in Atrial Fibrillation (SPAF) | 3 | 76% | 0 | 15 |
| Mesothelioma | 3 | 80% | 380 | 0 |
| Autoimmune Disorders | 3 | 61% | 50 | 0 |
| Kidney Transplant Rejection | 2 | 67% | 272 | 21 |
| Anemia Due to Chronic Kidney Disease | 2 | 80% | 123 | 55 |
| Bladder Cancer | 2 | 75% | 51 | 45 |
| COVID-19 Treatment | 2 | 65% | 24 | 0 |
| Cerebral Edema | 1 | 80% | 2 | 0 |
| Influenza | 1 | 65% | 0 | 132 |
| Chronic Cough | 1 | 90% | 44 | 0 |
| *Totals* | *242* | *-* | *11,022* | *3,615* |
| *Mean* | *8.64* | *73%* | *393.68* | *129.14* |
| *Median* | *6* | *71%* | *171* | *29* |
| *Std. Dev.* | *9.15* | *11%* | *628.6* | *219.12* |

To test whether an indication’s exposure to the Medicare-aged population predicts the size of small molecule investments, we utilized a multiple regression of the natural log of investments by an indication’s exposure to the Medicare-aged population and a dummy variable for investments made before and after the passage of the IRA. The results indicated a statistically significant negative relationship, with an impact of roughly minus one-half on the size of investments for small molecules (p < 0.0018) (Figure 5).

**Figure 5.**
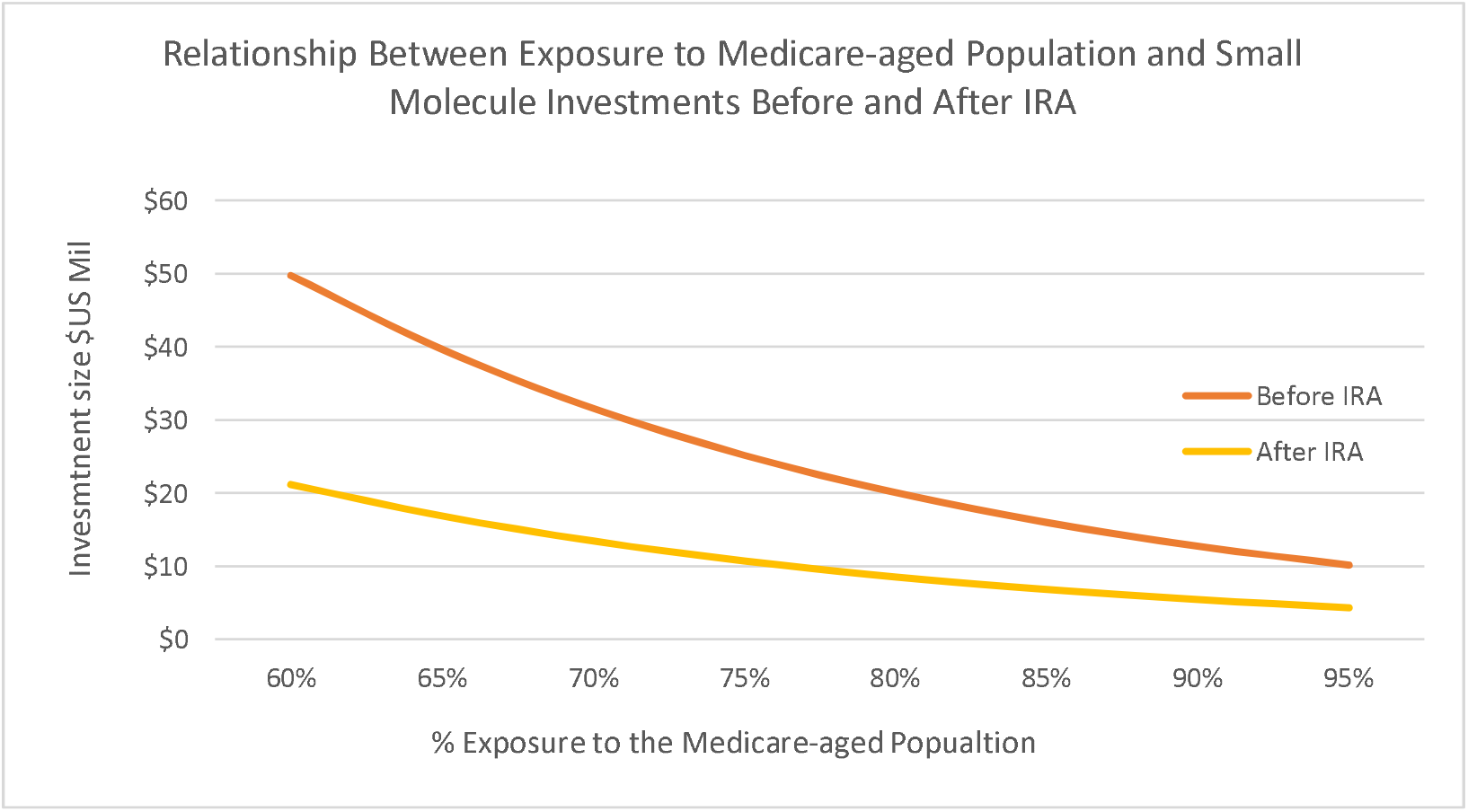
Estimated multiple regression relationship between small molecule investments and exposure to Medicare-aged population and time periods pre - and post-IRA. Data for 1/1/2018 – 8/16/2024 on U.S. companies ≤ $2billion valuation. Investments for phase I and II measured in 2024 constant dollars.

If this impact on small molecule investments is due to the general economic environment post-pandemic and not the IRA, as stated in the October 27, 2024, letter from Phillip L. Swagel, Director of the CBO (15), we would expect to observe similar results in large molecules. However, when we utilized a multiple regression of the natural log of large molecule investments by an indication’s exposure to the Medicare-aged population and a dummy variable for investments made before and after the passage of the IRA, the results showed no statistically significant difference in the median of large molecule investments before and after the IRA’s implementation.

The median frequency of an investment’s large molecule exposure to the Medicare-aged population is unchanged at 59% before and after the introduction of the IRA (Figure 6). We interpret this result to mean that, post-IRA, investors perceive large molecules to be a lower investment risk than small molecules.

**Figure 6.**
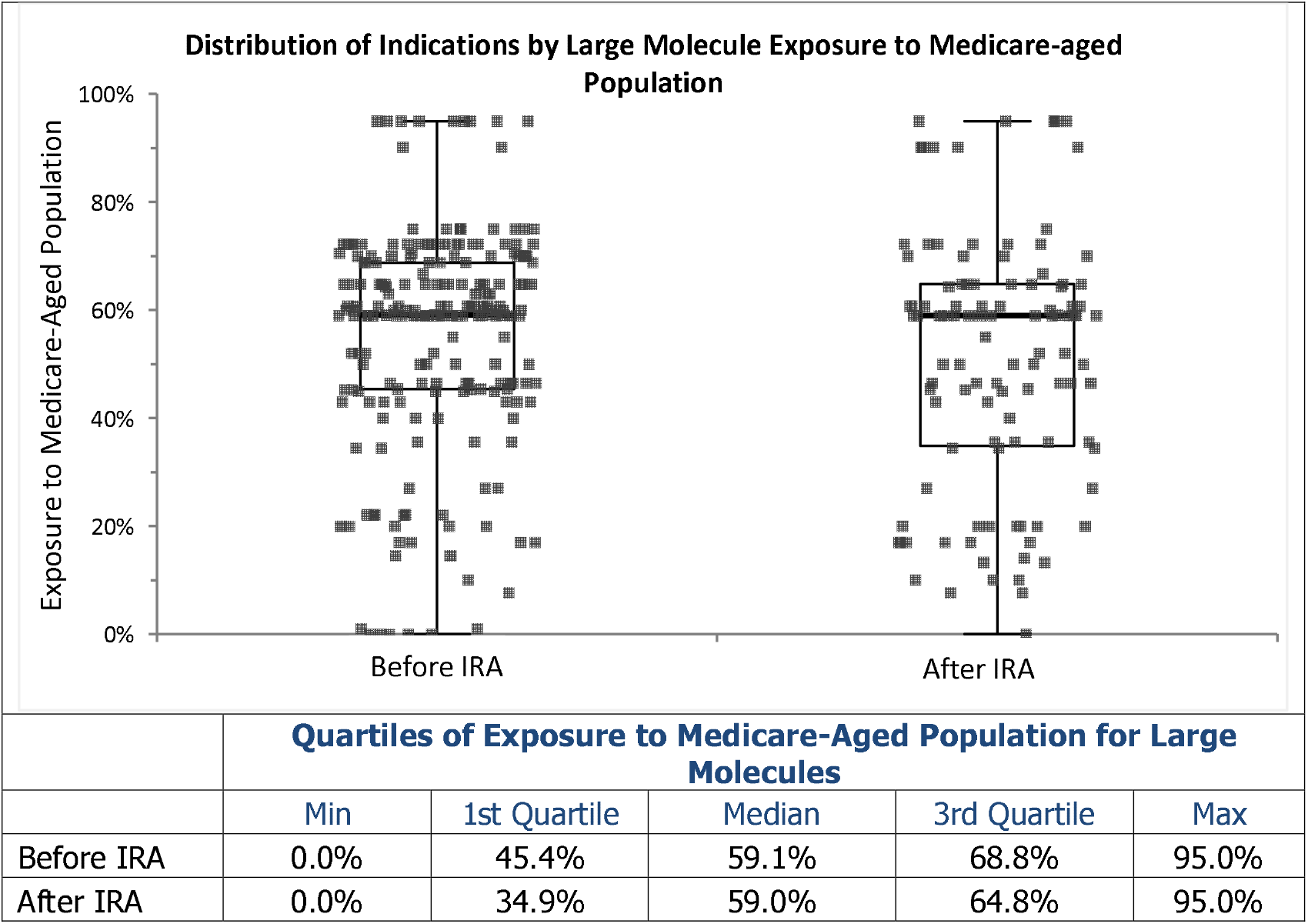
Dot plots show frequency of large molecule investments before and after the IRA vs their exposure to the Medicare-aged population. These indicate a decline in the number of large molecule investments post-IRA but no significant change in the nature of the distribution of such investments. Notably, as shown in the table, the frequency of such investments by quartiles of their exposure to the Medicare-aged population, pre- and post-IRA, is not statistically different. Investment data are for U.S. companies < $2 billion capitalization and for phase I and II trials from 1/1/2018 – 7/31/2024. Investment values in 2024 constant US dollars, N = 327.

## Discussion

The disconnect between the early- and late-stage IRA impacts has been discussed and debated, most notably between the CBO and the U.S. House of Representatives Budget Committee Health Care Task Force (16). Many of the previous studies claiming negligible impacts of the IRA have incorporated institutional large-scale financing of late-stage phase III therapies and have not explicitly investigated smaller companies targeting novel therapeutics and mechanisms of action reliant upon early-stage venture capital financing, initial public offerings, etc.

The analysis of this paper found, at the cohort level, that early-stage investors and small companies with assets under development consciously changed their behavior and investment activity in response to the altered financial risks caused explicitly by the IRA. As the average length of phase II and III clinical trials is roughly 40 months each, we expect these reductions to impact the rate of FDA approvals in 5 to 6 years (17).

The paper’s analysis also found evidence of a post-IRA decline in the size of investments into small molecules based upon an indication’s exposure to the Medicare-aged population. This suggests investors are now avoiding those indications with high exposure to the Medicare-aged population due to a change in the perceived risks of investing in large versus small molecule assets. In contrast, no such change was observed in the size and totality of investments into large molecules based on exposure to the Medicare-aged population.

Further support for the thesis of reduced investments in therapies with high exposure to the Medicare-aged population was the presence of three significant outlier investments into large molecules, each over $1 billion, undertaken after the passage of the IRA. For each of those investments, their value of their measured exposure to the Medicare-aged population was in the lowest quartile, i.e., among those investments with the lowest exposure to the Medicare-aged population. This highlights the lower relative risk and potentially higher return of large molecules compared to small molecules since, under the IRA, Medicare sets prices for large molecules in year 13 but in year 9 for small molecules, thereby enhancing revenue generation for large molecule relative to small molecule therapies.

## Limitations

According to Vogel et al., “In 2023, biopharmaceutical VC funds reportedly raised $21 billion in new capital, less than the $31 billion peak in 2021.” (18). Our research does not dispute that a downturn in the biopharmaceutical market may be impacting our results. However, our study does demonstrate significant changes in investing behavior that are predictable based on the projected impacts of the IRA. Such changes are evident in the observed movements away from small molecule investments for indications with a high exposure to the Medicare-aged population, the increased size of aggregate investment into large molecules for indications with low exposure to the Medicare-aged population, as well as the unchanged frequency of investments into large molecules when compared to small molecules relative to their exposure to the Medicare-aged population.

## Conclusion

Prior research on the potential impacts of the IRA on the biopharmaceutical ecosystem has primarily focused on the aggregate (mean) impacts and failed to examine segmented investments by indication or the degree of an indication’s exposure to the Medicare-aged population. As a result, prior research has obfuscated and primarily overlooked the IRA’s impacts on early-stage investment and drug development behavior. As the development time for new therapies is roughly 10 years from an FDA Investigational New Drug application to approval (19), the evidence presented in this paper of an observed 35% decline in clinical trial launches for the Medicare-aged population and a 70% decline in funding for early-stage developments in small molecules indicates significant negative impacts on the population the IRA legislation is allegedly designed to aid, namely, the Medicare-aged population requiring effective new therapies in areas of high unmet medical need..

## Data Availability

All data produced are available online

https://1drv.ms/u/c/fd1ceff1664dae51/EcIqeDD0_IFBrjgr01xmSYwBRxPxlbcVtZiqTlUmxH6HzQ?e=zvoYEQ

